# Effects of COVID-19 2021 lockdown on the sleep quality and mental health of undergraduate medical and dental students of Pakistan: A cross-sectional study

**DOI:** 10.1101/2021.07.17.21260690

**Authors:** Naveen Siddique Sheikh, Aiza Anwar, Iqra Pervaiz, Zunaira Arshad, Huma Saeed Khan, Farida Hafeez

**Affiliations:** CMH Lahore Medical College and Institute of Dentistry, Lahore, Pakistan

**Author notes:** Corresponding author: Dr. Naveen Siddique Sheikh.

**Keywords:** Anxiety, COVID-19, depression, sleep, medical students, mental health, Pakistan, Patient Health Questionnaire, Pittsburgh Sleep Quality Index

## Abstract

**Background and objectives:** The COVID-19 pandemic has been recognized as a cause of direct and indirect psychological consequences that impact mental health such as acute stress disorders, anxiety, irritability, poor concentration, and insomnia. This study was planned to evaluate the sleep quality and mental health of undergraduate students amidst the COVID-19 lockdown of 2021.

**Materials and Methods:** This observational cross-sectional study was conducted in Lahore, Pakistan, where 261 undergraduate medical and dental students enrolled at a private medical and dental school were approached from March to May 2021. The Pittsburg Sleep Quality Index (PSQI) was used to identify the sleep quality along with the Generalized Anxiety Disorder Scale (GAD-7) to establish anxiety symptoms and the Physical Health Questionnaire (PHQ-9) for depression symptoms.

**Results:** The results show that 212 (81.2%) female and 49 (18.8%) male students participated in the study. Of the participants 75.1% experienced poor sleep quality, 90% had symptoms of depression, and 85.4% had symptoms of anxiety. The mean score on the PSQI scale was 8.59±4.10, on the GAD-7 scale was 11.36±5.94 and on the PHQ-9 scale was 13.70±6.81. Multiple regression analysis showed that anxiety symptoms (*β* = 0.315, p = 0.000) and depression symptoms (*β* = 0.398, p = 0.000) were significant predictors of sleep quality amongst the undergraduate medical and dental students.

**Conclusion:** A high majority of the study participants are experiencing poor sleep quality along with suffering from depression and anxiety amidst the COVID-19 lockdown. It is concluded from the analysis that anxiety and depression symptoms are significant predictors of sleep quality. Relevant authorities need to set up systems that help undergraduate medical students in alleviating and coping with these symptoms midst the COVID-19 pandemic.

## INTRODUCTION

The index case of the novel severe acute respiratory syndrome coronavirus 2 (SARS-CoV-2) was reported between mid-October to mid-November 2019(1). World Health Organisation (WHO) declared the disease spread a global health emergency on 31st January 2020, and on 11th March 2020 the coronavirus disease 2019 (COVID-19) was labeled a pandemic (2). Since then, as of 14th July 2021, there have been 187 million confirmed cases of COVID-19 infection and 4 million deaths due to the disease worldwide (3). COVID-19 caused a global GDP (gross domestic product) loss of 4.5% which correlates to an estimated economic loss of $3.94 trillion USD to the global economy (4). Subsequently, after being labeled a pandemic, strategies such as social distancing, quarantine, and masks in public places were announced to mitigate the spread of the infection. In Pakistan, the index case of COVID-19 was confirmed on 26^th^ February 2020, in Karachi city by the Ministry of Health, Government of Pakistan (5) and a National Action Plan for COVID-19 disease containment was announced by the Government of Pakistan on 12^th^ March 2020 (6) to curb the spread of the infection and by 23^rd^ March 2020, Pakistan went under its first national lockdown (7). Travel restrictions, mandatory school closures, and the suspension of non-essential commercial operations and industries were among the lockdown measures. To avoid becoming infected, people were advised to remain at home and keep themselves socially isolated. While lockdowns are unfavorable for individuals, they also have detrimental effects on the economy of a country, to help mitigate the economic loss, some essential industries were allowed to open up by mid-April but social gathering restrictions were not eased.

Previous outbreaks of Coronavirus such as SARS, MERS, and the Ebola outbreak have been linked to increased psychological distress among the affected population(8–10). Psychological responses to the outbreak included aberrant conduct, emotional and defensive behaviors along with reaction formation(11). The psychosocial impact of disease outbreaks appears to be significantly greater during quarantine measures. Social isolation and quarantine have been associated with anxiety, irritability, insomnia, panic, high-stress levels, and depression. Furthermore, quarantining is also linked to acute stress and trauma-related diseases, especially in high-risk groups like health workers(12). Isolation, restlessness, frustrations, fear of getting infected, a loss of autonomy, and worries regarding family/friends are just a few of the elements that might impair mental health during a quarantine. During past pandemics, poor sleep quality and elevated psychological distress were also well-established (13). Poor sleep, specifically, was linked to negative emotions, symptoms of depression, and a high risk of mental health issue development(14) The effect of the pandemic was also felt by the education system as everything shifted to online remote learning bringing about new challenges due to a lack of skill for operating technological resources(15). This was particularly of concern for students in the medical field as the complete shift to E-learning hindered their attainment of practical knowledge and experiences(16). A pandemic coupled with a drastic shift in learning only heightened the pressure and stress felt by the medical students, inadvertently affecting their sleep quality and mental health(13).

To our knowledge, no study has been done in Pakistan pertaining to the effect of the COVID-19 pandemic on both, the sleep quality and mental health of undergraduate medical students. In light of the lack of literature on this key issue, the current study was planned to establish the effect of the COVID-19 outbreak on the sleep quality and mental health of undergraduate medical and dental students at a tertiary care teaching institute in Lahore, Pakistan.

## METHODOLOGY

This observational cross-sectional study was conducted from March to May 2021 at a teaching institute affiliated with a tertiary care hospital in Lahore Pakistan, after obtaining approval from the Institution’s ethical review board. A non-probability consecutive sampling technique was used. All medical and dental students enrolled in the institute were invited to participate in the study. The sample size was calculated by using the Raosoft sample size calculator by keeping a 5% margin of error, 95% confidence interval, and 1050 as the population size with 50% response distribution. The sample size calculated was 260. Students with prediagnosed psychiatric illness and insomnia were excluded from the study. A research questionnaire constructed on google-forms was disseminated via online social media platforms (Facebook, WhatsApp, and Gmail) amongst the college students. One response per person was allowed to be submitted through response setting restrictions. Informed electronic consent was the first part of the questionnaire. Participants were explained the voluntary nature of their participation, thereafter their consent was sought prior to filling out the questionnaire and collecting data. The second part collected information regarding demographic variables i.e age, gender, year of enrollment, and bedtime.

The last part comprised of the Patient Health Questionnaire-9 (PHQ-9) (17) to measure depression symptoms, Generalized Anxiety Disorder-7 (GAD-7) (18) to measure anxiety symptoms, and Pittsburgh Sleep Quality Index (PSQI)(19) to measure sleep disturbances and quality fo sleep over the past month. Data were analyzed via SPSS (Statistical Package for the Social Science) software version 26. Frequency and percentage were calculated for qualitative variables i.e age groups, gender, year of enrollment, and course of enrollment. Mean ± SD was calculated for quantitative variables i.e duration of sleep, and PSQI, GAD-7, and PHQ-9 scores. Multiple regression analysis was done to evaluate the effect of independent variables (age, gender, year of enrollment, course of enrollment, PHQ-9 score, GAD-7 score) on the PSQI score (dependent variable). A *p*-value of < 0.05 was considered statistically significant.

## RESULTS

A total of 261 undergraduate medical and dental students participated in the study. Of the respondents, 212 (81.2%) were females. A broad score cut-off value of greater than and equal to four (≥4) on the GAD-7 and PHQ-9 scales indicated the presence of anxiety and depression symptoms respectively. A broad score cut-off value greater than five (>5) on the PSQI scale differentiated between good sleepers and poor sleepers. Poor sleepers amounted to 75.1% of the participants, 90% of the participants had symptoms of depression, and 85.4% of participants had symptoms of anxiety as reflected in **Table 1** of participant demographics.

**Table 1:**
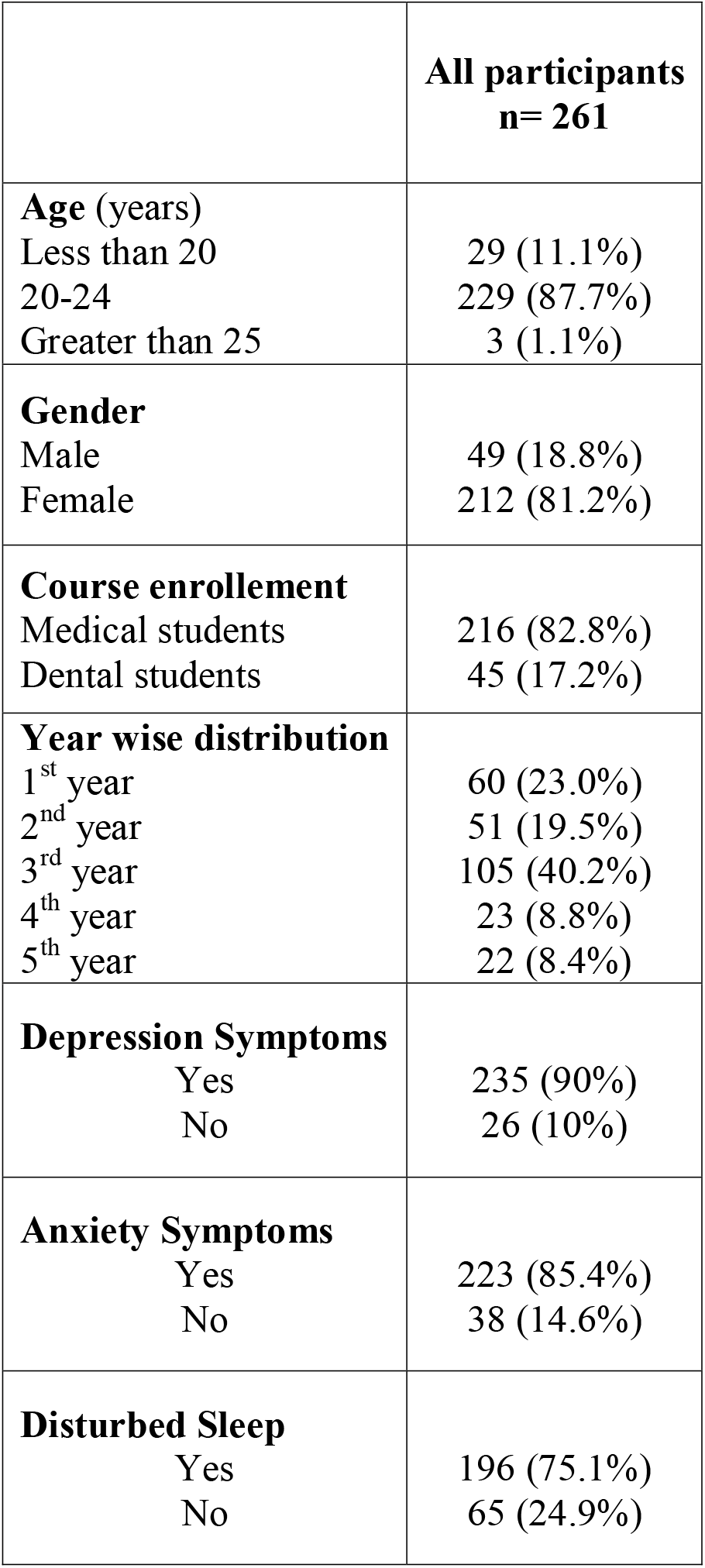
Participant demographics and classification according to scale cut-offs.

|  |  |
| --- | --- |
|  | **All participants**<br>**n= 261** |
| **Age (years)**<br>Less than 20<br>20-24<br>Greater than 25 | 29 (11.1%)<br>229 (87.7%)<br>3 (1.1%) |
| **Gender**<br>Male<br>Female | 49 (18.8%)<br>212 (81.2%) |
| **Course enrollement**<br>Medical students<br>Dental students | 216 (82.8%)<br>45 (17.2%) |
| **Year wise distribution**<br>1<sup>st</sup> year<br>2<sup>nd</sup> year<br>3<sup>rd</sup> year<br>4<sup>th</sup> year<br>5<sup>th</sup> year | 60 (23.0%)<br>51 (19.5%)<br>105 (40.2%)<br>23 (8.8%)<br>22 (8.4%) |
| **Depression Symptoms**<br>Yes<br>No | 235 (90%)<br>26 (10%) |
| **Anxiety Symptoms**<br>Yes<br>No | 223 (85.4%)<br>38 (14.6%) |
| **Disturbed Sleep**<br>Yes<br>No | 196 (75.1%)<br>65 (24.9%) |

The scores computed from the three scales were cut to define groups having minimal, mild, moderate, and severe symptoms of anxiety, depression, and sleep disturbances. PSQI score categories were defined on score cut-off values as Minimal (≤5) Mild (6-8) Moderate (9-12) Severe (>12) for sleep disturbances and quality of sleep in the preceding month. PHQ-9 score categories were formulated on score cut-offs as Minimal (1-4) Mild (5-9) Moderate (10-14) Severe (15-27) reflecting the different groups of depression symptoms. GAD-7 score categories were defined by score cut-off values of Minimal (0-4) Mild (5-9) Moderate (10-14) Severe (15-21) for anxiety symptoms. **Table 2** elaborates on the percentage of participants in each score classification along with score range, mean and standard deviation.

**Table 2:** Score details for depression, anxiety, and sleep scales.

| Number of participants n=261 | PSQI | PHQ-9 | GAD-7 |
| --- | --- | --- | --- |
| Mean±SD | 8.59±4.10 | 13.70±6.81 | 11.36±5.94 |
| Range | 1-20 | 0-27 | 0-21 |
| **Score categories** |  |  |  |
| Minimal | 65 (24.9%) | 26 (10.0%) | 38 (14.6%) |
| Mild | 70 (26.8%) | 52 (19.9%) | 64 (24.5%) |
| Moderate | 77 (29.5%) | 61 (23.4%) | 75 (28.7%) |
| Severe | 49 (18.8%) | 122 (46.7%) | 84 (32.2%) |
| PSQI: Pittsburgh sleep quality index<br>PHQ: Patient health questionnaire<br>GAD: Generalised anxiety disorder |  |  |  |

The PSQI scale comprised of seven components which were computed together to get a Global PSQI score for each participant. The Global PSQI score ranges from 0 to 21, where a score greater than five reflects poor sleep quality. Of the participants, only 65 (24.9%) individuals reported being good sleepers. The average sleep duration of the participants was 7.80 ± 2 hours. On average it took 31.98 minutes to initiate sleep for the participants, a total of 68 participants (26.1%) reported using a hypnotic once or twice a week to induce sleep, and 34 participants (13.0%) reported using a hypnotic three or more times in the past month to help them sleep.

**Table 3** reflects the percentage-wise distribution of participants for various responses to the PSQI questionnaire showing sleep disturbance.

**Table 3:** Pittsburgh sleep quality index (PSQI); sleep disturbance detail of the participants.

| Variables | Total Sample (n=261) |
| --- | --- |
| Sleep duration (hours) ± SD | 7.80 ± 2.00 |
| >7 hours | 174 (66.6%) |
| 6-7 hours | 25 (9.6%) |
| 5-6 hours | 32 (12.3%) |
| < 5 hours | 30 (11.5%) |
| Sleep latency (minutes) ± SD | 31.98 ± 28.34 |
| Difficulty initiating sleep |  |
| Yes | 175 (67%) |
| No | 86 (33.0%) |
| Difficulty maintaining sleep/early morning awakening |  |
| Yes | 184 (70.5%) |
| No | 77 (29.5%) |
| Had nightmares |  |
| Yes | 168 (4.4%) |
| No | 93 (35.%) |
| Used hypnotics to induce sleep |  |
| Yes | 211 (80.8%) |
| No | 50 (19.2%) |

The PSQI scale component mean ± SD for the seven components is detailed in **Table 4**. The mean global score obtained by the male participants on the PSQI scale was 7.87 ± 4.12 and the mean global score for females was 8.76 ± 4.09.

**Table 4:** Pittsburgh sleep quality index (PSQI) seven component analysis.

| Component | Mean | Standard deviation |
| --- | --- | --- |
| 1. Sleep duration | 0.68 | 1.07 |
| 2. Sleep disturbance | 1.29 | 0.62 |
| 3. Sleep latency | 1.65 | 1.14 |
| 4. Daytime dysfunction due to sleepiness | 0.70 | 0.81 |
| 5. Sleep efficiency | 1.21 | 1.27 |
| 6. Overall sleep quality | 1.71 | 1.10 |
| 7. Sleep medication use | 1.32 | 0.93 |
| **Global PSQI score** | **8.59** | **4.10** |

Multiple linear regression was performed to find the association between socio-demographic variables, PHQ-9 scores, GAD-7 scores, and the PSQI scores of the participants. The model was significant and the independent variables predicted 45.1% of the variance in sleep quality (PSQI scores) of the participants. Analysis showed that the presence of anxiety symptoms significantly predicted the sleep quality of participants (*β* = 0.315, *p* = 0.000). Similarly, the presence of depression symptoms also significantly predicted the sleep quality of participants (*β* = 0.398, *p* = 0.000). **Table 5** shows the multiple regression analysis of the association of independent variables as predictors of sleep quality (PSQI scores) of the participants.

**Table 5:** Results of the multiple linear regression model analyzing associations with sleep quality (ΔR^2^=0.451). The unstandardized regression coefficients represent the change in the likelihood of having poor sleep quality for a one-unit increase in the predictor variable.

| Variables | B | SE | $\beta$ | 95% CI | p-Value |
| --- | --- | --- | --- | --- | --- |
| Age | -0.218 | 0.615 | -0.018 | -1.428 - 0.993 | 0.724 |
| Gender | 0.411 | 0.489 | 0.039 | -0.551 - 1.374 | 0.401 |
| Course enrollment | -0.634 | 0.507 | -0.058 | -1.633 - 0.365 | 0.212 |
| Year of enrollment | 0.036 | 0.175 | 0.010 | -0.309 - 0.381 | 0.836 |
| **GAD-7 score** | **0.217\*** | 0.053 | 0.315 | 0.113 - 0.322 | **0.000\*** |
| **PHQ-9 score** | **0.240\*** | 0.047 | 0.398 | 0.148 - 0.332 | **0.000\*** |

Note: Dependent variable: PSQI (Pittsburgh sleep quality index) scores
B = Unstandardized Regression Coefficient, $\beta$ = Standardized Regression Coefficient,
SE = Standard Error for Beta, CI = Confidence Interval, \* = $p \leq 0.05$
GAD-7 = Generalised anxiety disorder-7, PHQ-9 = Patient Healthcare Questionnaire-9.

## DISCUSSION

The results showed that the undergraduate medical and dental students had long sleep latency, difficulty initiating and maintaining sleep. In addition, the presence of anxiety and depression symptoms were significant predictors of poor sleep quality amongst the participants; with an increasing amount of symptoms, the sleep quality of the participants dropped. Among the participants from our study, 196 (75.1%) were poor sleepers, 235 (90%) had symptoms of depressions, and 223 (85.4%) had symptoms of anxiety, these numbers are significantly higher from the pooled prevalence of 34% for depression symptoms, 32% for prevalence of anxiety and 33% for prevalence of sleep disturbances amongst higher education students during the COVID-19 pandemic as reported by a meta-analysis(20).

The average PSQI score of the participants was 8.59 ± 4.10, which was much higher than 6.87 ± 3.71 a previous study done at the same teaching institute in Lahore in 2018, highlighting the adverse effect of the COVID-19 pandemic on the sleep quality of the undergraduate students (21). In our study, we did not find any significant association between the socio-demographic variables and sleep quality, this is supported by a study done in Wuhan which also did not find any significant association between socio-demographic variables and sleep quality (22), however, previous studies have suggested a significant association between female gender, second-year academic students and sleep disturbance with a reported increasing trend of sleep disturbance (23).

This research warrants enhancements and does not come without its limitations. The study is cross-sectional which hinders disentangling causality of whether the sleep disturbance is due to the COVID-19 pandemic directly or were there confounding factors that contributed to sleep disturbances as well. Though causality can not be determined, the study still, however, suggests that the pandemic might have an effect on sleep disturbance and the mental health of undergraduate medical and dental students. Even though the sample size is adequate, the results should be generalized for the entire population of undergraduate medical and dental students of Pakistan with caution.

## CONCLUSION

According to our study, a large proportion of undergraduate medical and dental students are found to be suffering from depression, have anxiety symptoms, and are experiencing poor sleep quality. A substantial amount of participants are also making use of hypnotics to induce sleep which reflects the toll the COVID-19 pandemic associated lockdown is taking on their mental health and sleep quality. These findings highlight the need for necessary intervention by respective authorities to help alleviate the symptoms experienced by the students and set up a proper social support system for the students in these unprecedented times of a pandemic.

## Data Availability

Data (SPSS file) can be requested from the corresponding author via email.

